# Vaccination of Front-Line Workers with the AstraZeneca COVID-19 Vaccine: Benefits in the Face of Increased Risk for Prothrombotic Thrombocytopenia

**DOI:** 10.1101/2021.04.11.21255138

**Authors:** Amin Adibi, Mohammad Mozafarihashjin, Mohsen Sadatsafavi

## Abstract

**Background:** In March 2021, a number of regulatory and advisory bodies around the world recommended against using the AstraZeneca COVID-19 vaccine in younger adults pending further review of the risk for vaccine-induced prothrombotic immune thrombocytopenia (VIPIT). As an example, we consider the Canadian province of British Columbia (BC) which halted its front-line workers vaccination program with the AstraZeneca vaccine. The province received an additional 246,700 doses of AstraZeneca vaccine in the weeks before April 11th, enough to provide the first dose of vaccine to all unvaccinated front-line workers. It is unclear whether the alternative, mRNA vaccines can be immediately made available to front-line workers.

**Methods:** We reviewed the latest available evidence and used compartmental modelling to *1)* compare the expected number of deaths due to COVID-19 and VIPIT under the scenarios of immediately continuing vaccination of front-line workers with the AstraZeneca vaccine or delaying it in favour of mRNA vaccines from a societal perspective, and *2)* compare the individual mortality risk of immediately receiving the AstraZeneca vaccine with waiting to receive an mRNA vaccine later from a personal perspective.

**Results:** We estimate that if British Columbia continues the front-line worker vaccination program with the AstraZeneca vaccine, we expect to see approximately 45,000 fewer cases of COVID-19, 800 fewer hospitalizations, 120 fewer COVID-related deaths, and 2,300 fewer cases of Long COVID from April 15th to October 1st, 2021, for an expected number of VIPIT-related deaths of 0.674 [95% CI 0.414-0.997]. In the same period and in areas of high transmission (*R*_0_=1.30), the projected excess risk of mortality due to COVID-19 and VIPIT was significantly higher in the delayed vaccination with mRNA vaccines scenario (3.5 to 4.5 times higher risk) than that of immediate vaccination with the AstraZeneca vaccine for those between 30 and 69 years of age. In areas with lower levels of transmission (*R*_0_=1.15), the projected excess risk of mortality was 1.8 to 3.4 times higher in the delayed vaccination with mRNA vaccines scenario for those between 30 and 69 years of age. For those under 30, immediate vaccination with the AstraZeneca vaccine posed a higher risk than delayed vaccination with an mRNA vaccine, regardless of the level of transmission in the community.

**Conclusions:** The benefits of continuing immunization of front-line workers with the AstraZeneca vaccine far outweigh the risk both at a societal level and at a personal risk level for those over 40, and those over 30 in high-risk areas.

## 1. Background

On March 29th, 2021, Canada’s National Advisory Committee on Immunization (NACI) recommended against using the AstraZeneca (AZ) COVID-19 Vaccine for Canadians under the age of 55, due to concerns about vaccine-induced prothrombotic immune thrombocytopenia (VIPIT) based on European reports (NACI 2021; Greinacher et al. 2021; Schultz et al. 2021). On March 18th, 2021, the European Medicines Agency estimated the incidence of VIPIT at approximately 1 per 1,000,000 people vaccinated with the AZ vaccine (EMA 2021). A higher estimated rate of 1 per 100,000 by the Paul-Ehrlich Institut in Germany was published on March 19th (PEI 2021). It was this higher rate reported by the Paul-Ehrlich Institut (PEI) that led NACI to recommend against using this vaccine in adults under 55 years old (NACI 2021).

On April 1st, the UK Medicines & Healthcare Products Regulatory Agency (MHRA) updated its own previously reported data to report a total of 22 cerebral venous sinus thrombosis (CVST) and 8 other clot-related events from 18.1 million doses of the AZ vaccine (total incidence rate 1 in 600,000) (MHRA 2021a). On April 7th, MHRA concluded a possible link between the AZ vaccine and extremely rare clotting events and updated its data to report 79 UK cases of VIPIT (51 in women and 28 in men, all of them between 18 to 79 years old), including 44 cases of CVST and 35 cases of thrombosis in other major veins (incidence rate 1 in 250,000)(MHRA 2021b).

On the same day, the Pharmacovigilance Risk Assessment Committee (PRAC) of the European Medicines Agency (EMA) concluded that VIPIT should be listed as a very rare side effect of the AZ vaccine. PRAC noted that as of March 22nd, a total of 86 cases of VIPIT (62 cases of CVST and 24 cases of splanchnic vein thrombosis) and 18 fatalities out of about 25 million vaccine doses were reported in EudraVigilance, the EU drug safety database (EMA 2021). As of April 4th, 2021, 222 cases of VIPIT (169 cases of CVST and 53 cases of splanchnic vein thrombosis) had been reported to EudraVigilance out of around 34 million people who had received the AZ vaccine (EMA 2021).

BC had initially slated the AZ vaccine for outbreak control and front-line workers vaccination program. On March 29th and following NACI’s recommendation, BC paused using the AZ vaccine for those under 60 and put the front-line workers vaccination program on hold.

Canadian provinces received 1.5 million doses of the AZ vaccine from the US and another 316,800 doses from the COVAX program in between the two weeks period ending on April 11th (Government of Canada 2021). British Columbia received 246,700 doses from these two AZ deliveries, enough to finish providing the first dose to all remaining front-line workers.

The 300,690 doses of Pfizer-BioNTech and 105,900 doses of Moderna vaccines expected within the same time frame are currently allocated for the priority groups, indigenous population, and the currently ongoing age-based vaccination campaign. The AZ vaccine was initially allocated to front-line workers due to its easier handling and storage requirements. If it is not logistically possible to switch the vaccine allocation for above 55 years old age groups to the AZ vaccine and use either Pfizer-BioNTech or Moderna vaccines for younger front-line workers without delay, one might ask whether the benefits of immediately deploying the AZ vaccine for front-line workers outweigh the rare but serious risk for VIPIT.

Here, we provide a preliminary harm-benefit analysis of immediate vaccination of all front-line workers with the AZ COVID-19 vaccine. We based our analysis on mortality alone, and explore the risk both from a societal and individual risk perspective.

## 2. Methods

We assumed that BC allocates all 246,700 doses to front-line workers and that there is enough uptake that BC is able to administer all these doses. We compared immediately prioritizing front-line workers for the AZ vaccine (*Scenario A*) and asking them to wait to receive mRNA vaccines as part of the current age-based program (*Scenario B*). For harm-benefit analysis from a societal perspective, we compared expected number of deaths under each vaccination strategy.

We estimated the expected number of deaths due to VIPIT as *E*(*death*)_*VIPIT*_ = *d ×P* (*VIPIT*|*vaccine*) *× P* (*death*|*VIPIT*), where *d* is the number of doses administered, *P* (*VIPIT|vaccine*) is the risk of VIPIT after receiving each dose of the AZ vaccine, and *P* (*death* |*VIPIT*) is the case fatality for VIPIT. We did a probabilistic analysis in which appropriate probability distributions were assigned to model parameters for which relevant data were available.

We assumed that each dose of the vaccine is independently associated with the risk for VIPIT and that the risk of VIPIT is uniform across all age groups. The most recent estimate for these probabilities by EMA, and the estimates NACI used in its calculations are summarized in Table 1.

**Table 1.**
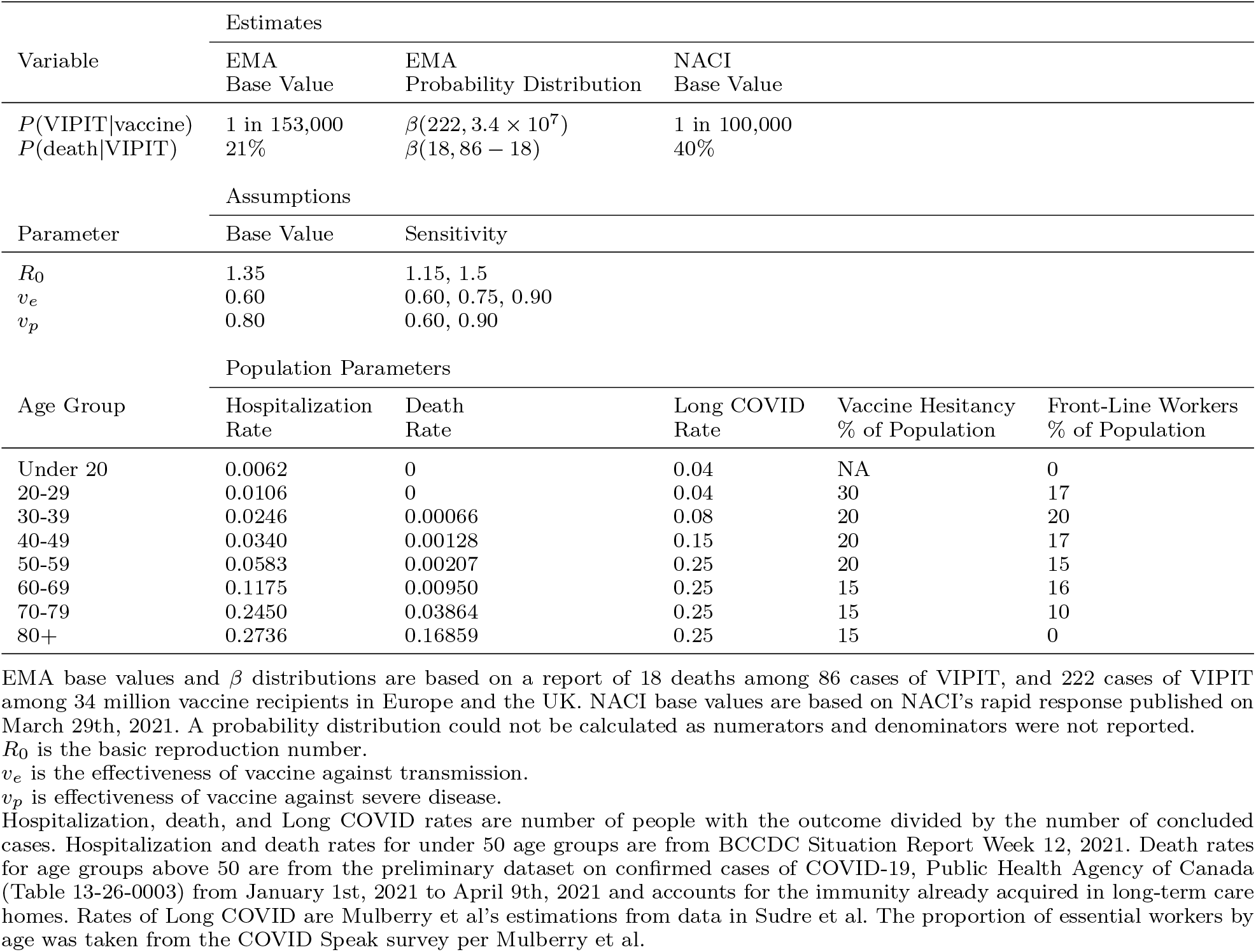
Harm-benefit parameters and model assumptions

| Variable | Estimates |  |  |  |  |
| --- | --- | --- | --- | --- | --- |
|  | EMA<br>Base Value | EMA<br>Probability Distribution | NACI<br>Base Value |  |  |
| $P(\text{VIPIT}\vert \text{vaccine})$ | 1 in 153,000 | $\beta(222, 3.4 \times 10^7)$ | 1 in 100,000 | | |
| $P(\text{death}\vert \text{VIPIT})$ | 21% | $\beta(18, 86 - 18)$ | 40% | | |
| Parameter | Assumptions |  |  |  |  |
|  | Base Value | Sensitivity |  |  |  |
| $R_0$ | 1.35 | 1.15, 1.5 | | | |
| $v_e$ | 0.60 | 0.60, 0.75, 0.90 | | | |
| $v_p$ | 0.80 | 0.60, 0.90 | | | |
| Age Group | Population Parameters |  |  |  |  |
|  | Hospitalization<br>Rate | Death<br>Rate | Long COVID<br>Rate | Vaccine Hesitancy<br>% of Population | Front-Line Workers<br>% of Population |
| Under 20 | 0.0062 | 0 | 0.04 | NA | 0 |
| 20-29 | 0.0106 | 0 | 0.04 | 30 | 17 |
| 30-39 | 0.0246 | 0.00066 | 0.08 | 20 | 20 |
| 40-49 | 0.0340 | 0.00128 | 0.15 | 20 | 17 |
| 50-59 | 0.0583 | 0.00207 | 0.25 | 20 | 15 |
| 60-69 | 0.1175 | 0.00950 | 0.25 | 15 | 16 |
| 70-79 | 0.2450 | 0.03864 | 0.25 | 15 | 10 |
| 80+ | 0.2736 | 0.16859 | 0.25 | 15 | 0 |

EMA base values and $\beta$ distributions are based on a report of 18 deaths among 86 cases of VIPIT, and 222 cases of VIPIT among 34 million vaccine recipients in Europe and the UK. NACI base values are based on NACI's rapid response published on March 29th, 2021. A probability distribution could not be calculated as numerators and denominators were not reported.
$R_0$ is the basic reproduction number.
$v_e$ is the effectiveness of vaccine against transmission.
$v_p$ is effectiveness of vaccine against severe disease.

We estimated the benefits of the AZ COVID-19 vaccine using a BC-specific age-structured COVID-19 compartmental model by Mulberry and colleagues that takes into account transmission, age-based contact structure, front-line worker status, and rising *R*_0_ due to variants of concern (Mulberry et al. 2021). The model included susceptible, exposed, infectious and recovered (SEIR) status and was based on the transmission model by Bubar et al (Bubar et al. 2021).

We ran the model from January 2021 to September 2021, which is when the vaccination campaign is expected to conclude. To follow BC vaccination strategy and case counts in the first three months of 2021, we held *R*_0_ at 1.03 from January 1, 2021 for 70 days during which people over 80 years old were eligible for vaccination. Age groups that were offered vaccination were considered to be vaccinated at a steady pace until everyone who is not vaccine-hesitant is vaccinated. Around the end of March, we raised *R*_0_ to either 1.15 or 1.35 to account for variants of concern gaining a foothold in BC and increased the pace of the vaccination program. We validated these assumptions by comparing model projections under these values against observed case counts for the January 1st to April 15th, 2021 period.

We assumed the first dose of the vaccine, regardless of the manufacturer, to be, on average, 80% effective against severe COVID-19 and 60% effective in preventing transmission. We further assume that all British Columbians will be offered a first dose of one of the approved COVID-19 vaccines before July 1st, 2021, and a second dose before the end of September 2021. We assumed the second dose to have no effect other than prolonging the immunity acquired after the first dose and posing a risk for VIPIT again. We did not consider the risk for anaphylaxis, as all vaccines seem to have a similar risk in that regard and the risk can be mitigated in the vaccination clinic. Population estimates that were used in the model are summarized in Table 1. COVID-19 age-based case fatality and hospitalization rates were obtained from BC Centre for Disease Control (BCCDC) (BCCDC 2021) and Public Health Agency of Canada (Table: 13-26-0003)(Statistics Canada 2021). For seniors, we used case fatality rates from January 1st, 2021 to April 9th, 2021 to account for the immunity already acquired in long-term care homes. Rates of Long COVID are Mulberry et al’s estimations from data in Sudre et al(Sudre et al. 2021). The proportion of essential workers by age was taken from the COVID Speak survey per Mulberry et al (Mulberry et al. 2021). Population of BC in each group was obtained from Statistics Canada (Statistics Canada 2017).

For harm-benefit analysis at an individual risk level, we weighed the probability of VIPIT-related death or *P* (death)_VIPIT_ = *P* (vaccine) *× P* (VIPIT|vaccine) *× P* (death|VIPIT), against the average probability of contracting COVID-19 and dying from it in each age groups, or *P* (death)_delayed vaccination_ = *P* (COVID-19) *×P* (death |COVID-19), where *P* (vaccine) is the probability of getting the AZ vaccine (assumed to be 1 here), and *P* (COVID-19) is the average probability of contracting COVID-19 due to delayed vaccination from April 1st, 2021 to July 1st, 2020.

We used results from our compartmental model to project mortality risk from COVID-19 due to delayed vaccination, and conducted sensitivity analyses around model assumptions as outlined in Table 1.

All the analysis was performed using publicly-available data and code. This manuscript is produced by an open-source and reproducible R Markdown script, which is available on Github.

## 3. Results

### 3.1. Model validation

Predicted epidemiological curve and age-stratified case counts showed good agreement with observed counts reported by BC CDC, except for the 80 and above age category where the model underestimated case counts (Figure 1), however, this age group had almost no implications on the compared scenarios.

**Figure 1.**
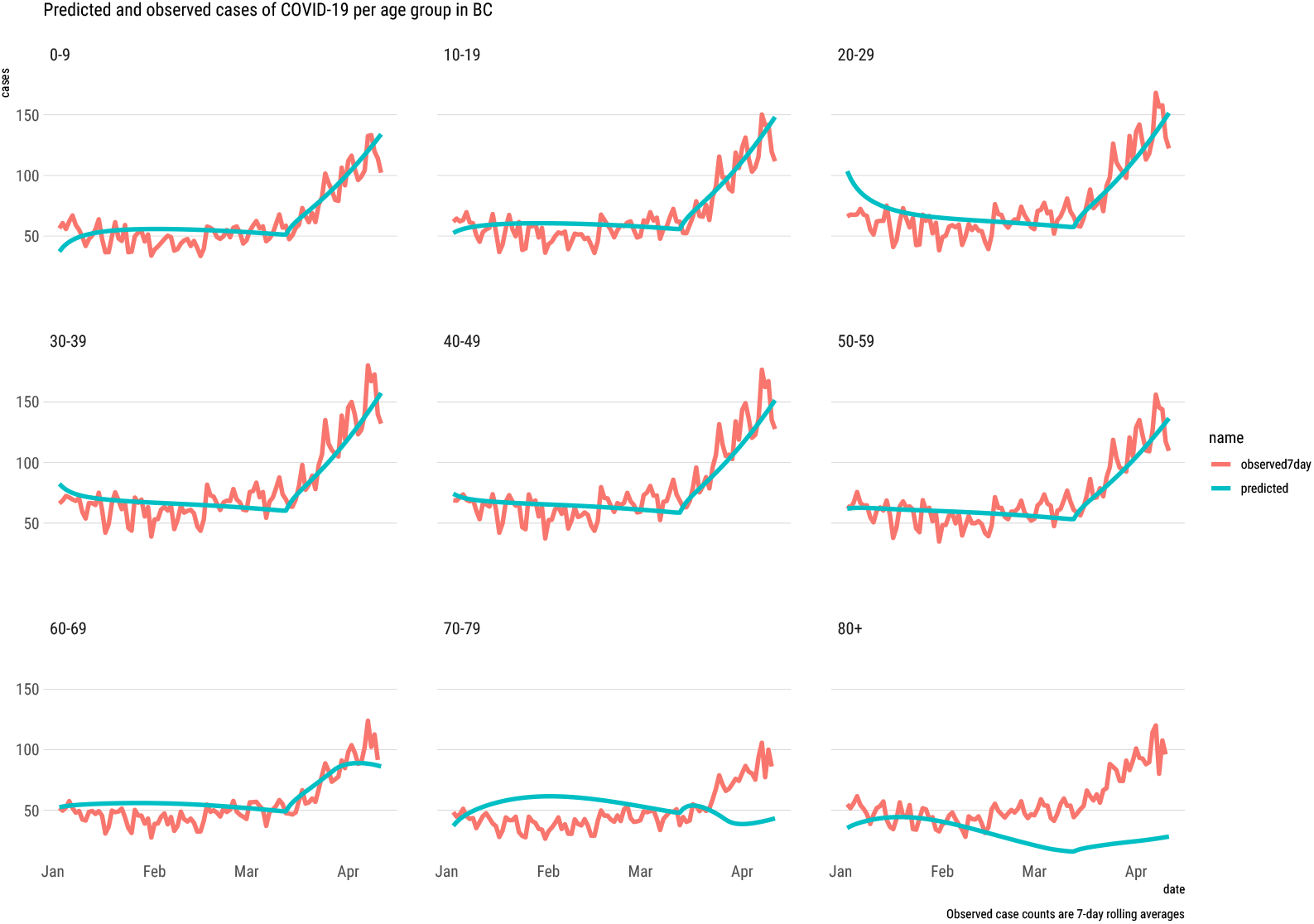
Face validity of model case counts

### 3.2. Harm-benefit from a societal perspective

EMA evidence as of April 4th, 2021 suggests that if we immediately offer a first dose of the AZ vaccine to all eligible front-line workers in BC, the expected number of VIPIT-related deaths by the end of June 2021 is 0.337 [95% CI 0.206-0.496], which means the probability of observing at least one VIPIT-related death in the same period is 28.6%. Adding the risk from the second dose, the expected number of VIPIT-related deaths until the end of summer is 0.673 [95% CI 0.412-0.997]. The probability of observing at least one VIPIT-related death till the end of the summer will be 49%.

NACI had based its analysis on the PEI estimates of a chance of 1 in 100,000 for VIPIT, and a mortality probability of 40%, based on the data that was available in late March. In this worst-case scenario, the expected number of deaths in BC would be 1 after the first dose is completed for all front-line workers and 2 after the second doses are delivered. Details of the calculations can be found in Appendix A.

Figure 2 shows the progression of the vaccination campaign, as well as projections for COVID-19 cases, hospitalizations, and deaths under the two scenarios of immediately prioritizing front-line workers (A) and the current scenario of asking them to wait to receive mRNA vaccines as part of the current age-based vaccination program (B).

**Figure 2.**
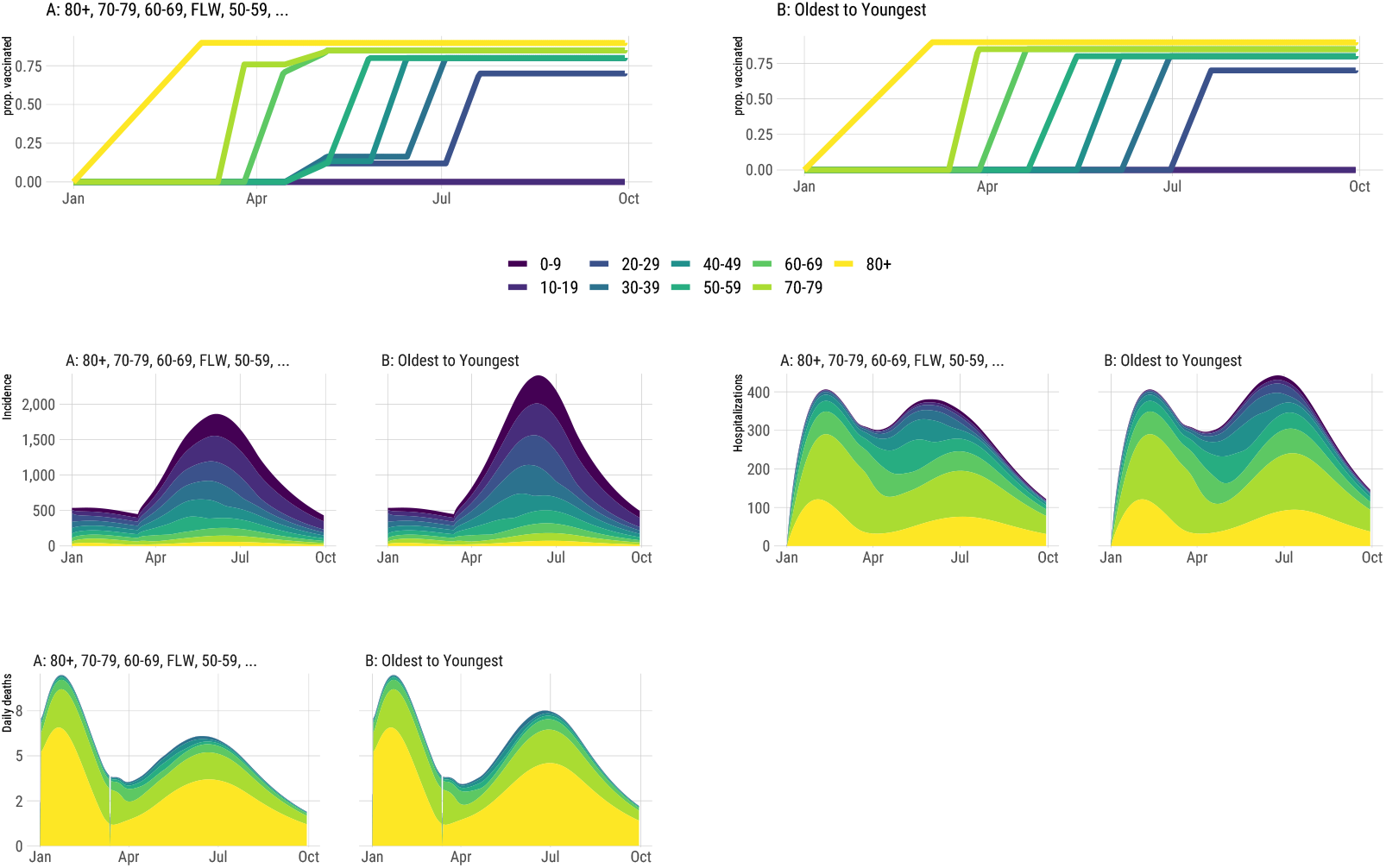
Top Panel: Projection of the progression of the vaccination for different age groups and front-line workers (FLW). Bottom Panel: Projection of COVID-19 cases, hospitalizations, and deaths from January 1st to October 1st, 2021.

In our analysis, Scenario A led to 45755 fewer cases of COVID-19, 811 fewer hospitalizations, 127 fewer deaths, and 2321 fewer cases of Long COVID, assuming *R*_0_ = 1.35. Appendix B includes results of the sensitivity analysis for a wider range of values for *R*_0_ and the effectiveness of the vaccine against transmission, *v*_*e*_.

### 3.3. Harm-benefit from an individual risk perspective

Figure 3 compares the risk of VIPIT-related mortality from 2 doses of the AZ vaccine and residual risk from COVID-19, with the mortality risk from COVID-19 due to delayed vaccination from April 1st to October 1st, 2021. We did the comparison under two scenarios of an *R*_0_ of 1.15 or 1.35, to represent different intensities for the third wave, or alternatively to represent different geographical parts of the province during the third wave. We calculated the mortality risk associated with VIPIT using both the latest and most comprehensive evidence by EMA, and the worst-case scenario using the evidence considered by NACI. Using EMA estimates, we found that under both *R*_0_ scenarios, the mortality risk due to COVID-19 to be much higher than the mortality risk associated with VIPIT in those over 40. Mortality risk from COVID-19 was also higher than that of VIPIT for the 30-39 age group, although the difference was negligible under *R*_0_ of 1.15 scenario. For the 20-29 age group, the estimated mortality risk of vaccination with the AZ vaccine was higher than that of COVID-19. Using the worst-case VIPIT estimates considered by NACI, mortality risk from COVID-19 was considerably higher than that of VIPIT for those over 60 in all areas and those over 30 in high-risk areas (*R*_0_ = 1.35).

**Figure 3.**
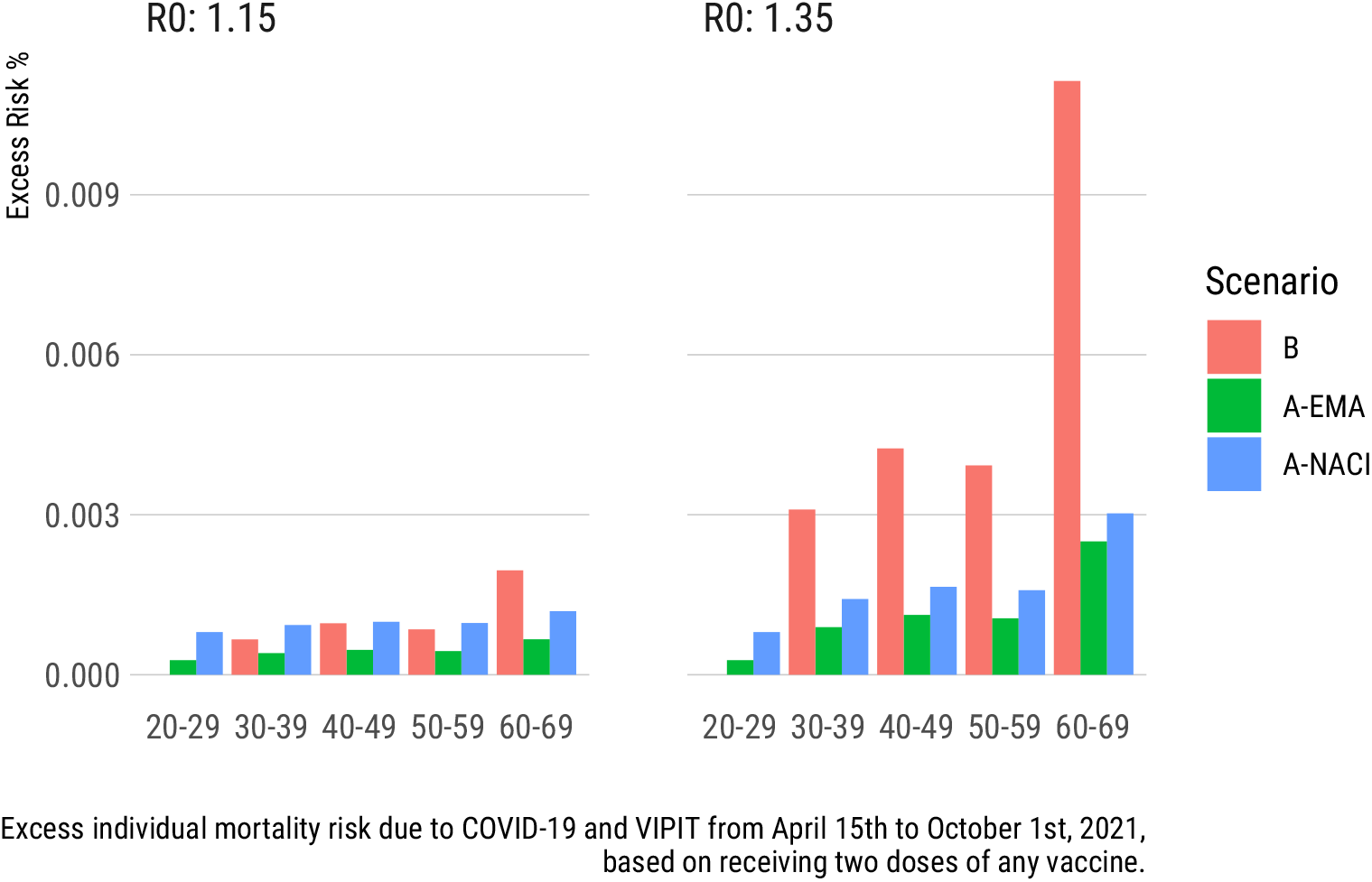
Comparison of excess mortality risk for different age groups based on the COVID-19 risk caused by delayed vaccination (B) and estimated residual COVID-19 and VIPIT risk (A) by the EMA and NACI

## 4. Discussion

In its analysis of AZ vaccine published on March 29th, 2021, NACI weighed the risk of adverse events against the age-stratified risk of mortality due to COVID-19, and concluded that the AZ vaccine should not be used in adults under 55 years of age pending an overall risk-assessment. Our analysis confirms that given the evidence available in late March (a risk of 1 in 100,000 for VIPIT and a 40% case fatality) and the lower rates of transmission (i.e. a lower *R*_0_) at that time, suspending the use of AZ vaccine in younger adults would have been advisable. However, as of April 8th, 2021, the evidence has evolved and the EMA is now reporting a risk of 1 in 153,000 for VIPIT and a 20% case fatality. These latest estimates together with a new wave of the disease have changed the harm-benefit landscape considerably. In addition, the benefits of the AZ vaccine go beyond preventing COVID-related mortality and include protection against other possible COVID-19 complications in younger adults including hospitalizations and associated risk of venous thromboembolism (VTE; Rates are about 2 folds higher in hospitalized COVID-19 patients than that of medical non-COVID-19 inpatients (Alberta Health Services 2021)), and Long COVID, as well as preventing onward transmission of the virus, as as suggested by the recent sharp decline of COVID-19 cases in the UK (GOV.uk 2021) and possible emerging signal in a recent observational healthcare worker/household study (Shah et al. 2021).

The UK vaccination program started on December 8th with the Pfizer-BioNTech vaccine and was complemented with the AZ vaccine since January 4th. The number of confirmed daily COVID-19 cases in the UK has plummeted from about 60,000 cases a day in early January 2021 when a national lockdown was imposed and about 3% of the population had received at least one vaccine dose, to about 11,000 cases per day on February 22, 2021 when a roadmap to easing the lockdown was announced, to about 6000 cases per day on March 8, 2021 when the first phase of easing public health restrictions was commenced (BBC 2021) and has continuously declined since then to just under 2500 cases as of April 14th, 2021, when 61.4% of the UK population had received one dose of a COVID-19 vaccine (GOV.uk 2021).

In addition to the death aversion resulting from strict public health measures, based on a recent analysis by Public Health England and the University of Warwick, it has been estimated that as of March 31, 2021, 10,400 deaths have been avoided in the UK solely due to the direct implementation of a nationwide vaccination program (indirect effects were not measured; 87.5% of these averted deaths were in the 80+ years old age group, 11.5% of them in 70-79, and 1% in 60-69 years old age group) (Public Health England 2021b). As about half of all vaccine doses administered in the UK have been AZ vaccines, and based on the estimated AZ vaccine efficacy of about 76% against symptomatic COVID-19 and 64% against any NAAT-positive COVID-19 infection between 22 and 90 days after the first dose (Voysey et al. 2021), and real-world single-dose AZ vaccine effectiveness of about 60% against symptomatic COVID-19 and 80% against COVID-19 hospitalization (Public Health England 2021a), it is clear that the AZ vaccine is effective in reducing the overall burden of COVID-19.

Potential prevention of onward transmission with the AZ vaccine could be especially critical for front-line workers during the current wave of COVID cases. Of note, two recent studies from Toronto, Ontario have shown that neighbourhoods with the highest proportion of front-line workers had per capita COVID-19 case and death rates that were 2.5-3 folds higher than that of neighbourhoods with the lowest share of front-line workers (Chagla et al. 2021, Rao et al. (2021)).

Based on our analysis, immediately making the AZ vaccine available to front-line workers is, assuming optimal uptake, net-beneficial by a wide margin from a societal perspective. Our analysis from an individual risk perspective shows that the risk of contracting COVID-19 and dying from it due to delayed vaccination is considerably higher than the risk of dying from VIPIT in those over 40, and also in those over 30 in high-risk areas.

For a public health intervention to be deemed ethically acceptable, being net-beneficial at a societal level is not enough in and of itself. Not all interventions that are net-beneficial at a societal level are net-beneficial for each member of the society, as those who carry the burden of the risk of adverse events may not be the same people who reap the benefits. We also recognize that many might intuitively consider mortality due a public health intervention in an otherwise healthy person to be ethically worse than failing to protect someone from mortality due to COVID-19. While we are not going to solve the trolley problem here, we believe that our conclusions hold regardless of the position we take with respect to the *doing vs. allowing harm* problem (Woollard and Howard-Snyder 2016), as long as the expected benefits outweigh the risk at a personal level, as seems to be the case for most age groups in our study.

Our analysis was based on the assumption that immediate deployment of alternative mRNA vaccines for front-line workers was not logistically feasible. If feasible, offering mRNA vaccines to front-line workers will be more in line with the principle of reciprocity outlined in the BC COVID-19 Ethical Decision-Making Framework (BC-CDC 2020), and the more general principle of justice in bioethics (McCormick and Min 2021), based on the fact that as of April 10th, 2021, no VIPIT cases have been linked to COVID-19 mRNA vaccines. From a vaccine efficacy point of view, determining the *superiority* of one vaccine to the other is not straightforward; there is more to the apparent but variable 4%-33% efficacy gap (symptomatic COVID-19; 7-14 days after second dose) between mRNA and AZ COVID-19 vaccines (Polack et al. 2020; Baden et al. 2021; AstraZeneca 2021; Emary et al. 2021) than meets the eye, including differences in study populations, settings, time periods (i.e. different incidence rates of COVID-19 from both non-variant of concern [VOC] and VOC SARS-CoV-2) at different times of the year), and vaccine storage requirements (Ledford 2021). Importantly, these vaccines were comparable in efficacy against severe COVID-19 in phase 3 trials (Abdool Karim and de Oliveira 2021; AstraZeneca 2021). Considering emerging real-world vaccine effectiveness data, two studies in the UK demonstrated that one dose of the Pfizer-BioNTech mRNA vaccine or the AZ vaccine have a comparable performance in terms of reducing rates of PCR-positive SARS-CoV-2 infection, symptomatic COVID-19, and COVID-19-related hospitalizations (Shrotri et al. 2021; Jamie Lopez Bernal et al. 2021).

On April 14th, 2021, Health Canada issued an advisory concluding that rare VIPIT events may be linked to the AZ COVID-19 vaccine and updated its label accordingly. Health Canada did not identify any specific risk factors and did not restrict the use of vaccine at this time (Health Canada 2021). Health Canada’s conclusions are consistent with our assumption that the risk of VIPIT is uniform across age groups and pave the way for Canadian provinces to make the AZ vaccine available to those under the age of 55. However, while the risk from VIPIT might be similar across age group, the risk from COVID-19 is not, and as such our age-based harm-benefit approach remains valid. Our findings are further corroborated by the the recent recommendation of the UK Joint Committee on Vaccination and Immunisation (JCVI) that the benefits of the AZ vaccine far outweigh the risk in 30 years old or older recipients (JCVI 2021).

## 5. Limitations

Our analysis from an individual risk perspective was based on average rates of COVID-19 and its related outcomes per age group. However, the true risk within age groups is still heterogeneous and is affected by many factors including but not limited to exposure, medical history, work environment, and socioeconomic status.

Our analysis did not consider social aspects of vaccine roll-out such as the effect of different roll-out strategies on uptake and vaccine hesitancy, as they were beyond our expertise. However, we recognize that each time a recommendation for vaccine safety is reversed, there might be a penalty in public trust which could fuel vaccine hesitancy. Potential for these effects should be weighed carefully by policy makers.

Our analysis is based on currently available estimated rates of 1 in million to 1 in 100,000 for VIPIT and might need correction should higher rates of this complication be reported.

We have not considered potential sex differences in the risk for VIPIT. Although cases identified to date have been predominantly female, it remains unclear whether this was due to more females receiving the AZ vaccine or due to an intrinsic difference in risk.

## 6. Conclusions

Current evidence suggests that benefits of immediate prioritization of front-line workers for vaccination with the AZ vaccine far outweigh the risk, both at a societal and at a personal level for those over 40 years of age, and those over 30 years of age in high-risk areas. Ultimately, in dynamic situations like this where the evidence is uncertain and evolving, vaccine roll-out decisions are judgment calls that need to take a complex network of medical, epidemiological, ethical, logistical, and societal considerations into account.

## Data Availability

Data and model used for this study are publicly available.

https://github.com/aminadibi/astrazenecaVIPIT

## 7. Appendices

## Appendix A. Calculations for expected number of death

Assuming that BC allocates all 246,700 doses to front-line workers, we can estimate the expected number of deaths due to VIPIT, *E*(*death*)_*VIPIT*_, as shown below. To err on the side of caution, we assume that each dose of the vaccine is independently associated with the risk for VIPIT and that the risk of VIPIT is uniform across all age groups. We also assume that there is enough uptake that BC is able to administer all these doses.

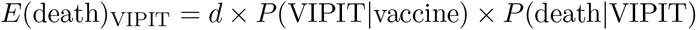

where d is the number of doses administered, *P* (VIPIT|vaccine) is the risk of VIPIT after receiving each dose, and *P* (death|VIPIT) is the case fatality for VIPIT.

According to the most recent data from UK and EU submitted to EudraVigilance (as of April 4th, 2021):

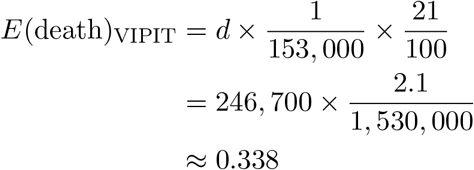

Considering both doses of the vaccine, we will have:

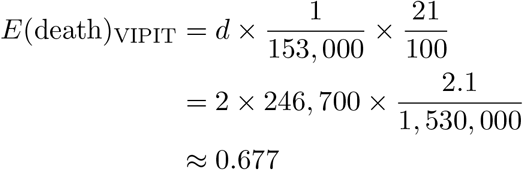

NACI had based its analysis on the more pessimistic estimates of a chance of 1 in 100,000 for VIPIT, and a mortality probability of 40%. In this worst-case scenario analysis, the expected number of deaths in BC would be 1:

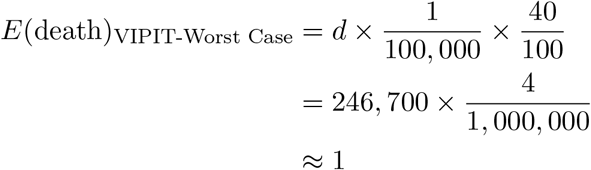

Considering both doses of the vaccine, the expected number of deaths in BC would be 2.

## Appendix B. Sensitivity Analysis

Figure B1 summarizes projected COVID-19 case counts, hospitalizations, and deaths, for a wider range of values for *R*_0_ and the effectiveness of the vaccine against transmission, *v*_*e*_. Sensitivity analysis on vaccine effectiveness against severe disease, *v*_*p*_, lead to similar results and conclusions with a slight variation in number of outcomes. As *v*_*p*_ increased, both the overall number of deaths and the number of deaths prevented decreased. Specifically, under *v*_*p*_ = 0.90, Scenario A led to 46779 fewer cases of COVID-19, 730 fewer hospitalizations, 102 fewer deaths, and 2137 fewer cases of Long COVID, assuming *R*_0_= 1.35. Figures B2 to B5 summarize main results when *v*_*p*_ = 0.90.

Under *v*_*p*_ = 0.60, Scenario A led to 43706 fewer cases of COVID-19, 910 fewer hospitalizations, 179 fewer deaths, and 2553 fewer cases of Long COVID, assuming R 0 = 1.35. Figures B6 to B9 summarize main results when *v*_*p*_ = 0.60.

**Figure B1.**
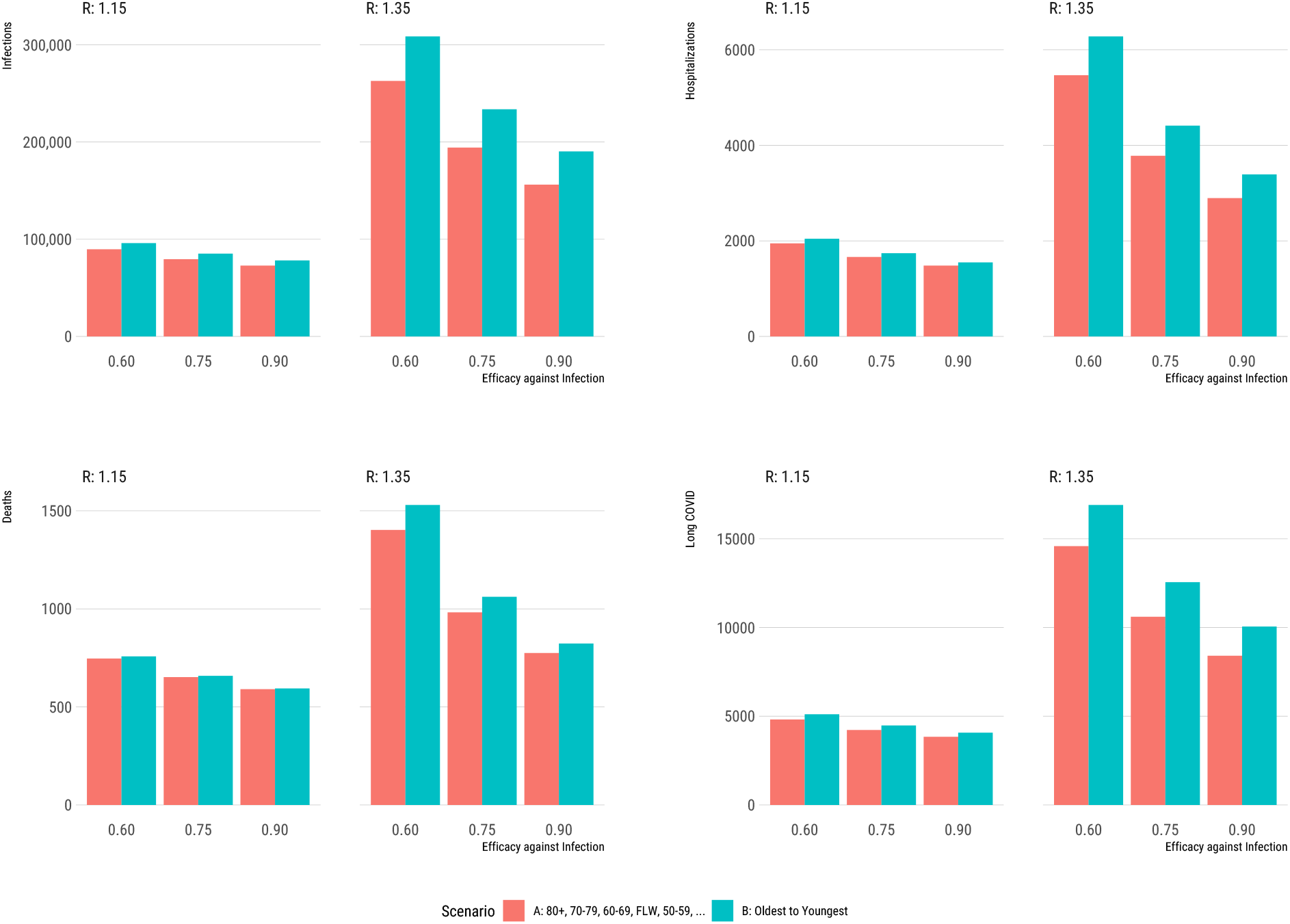
COVID-19 outcomes under different vaccination scenarios for different age groups and front-line workers (FLW) when *v*_*p*_ = 0.80

**Figure B2.**
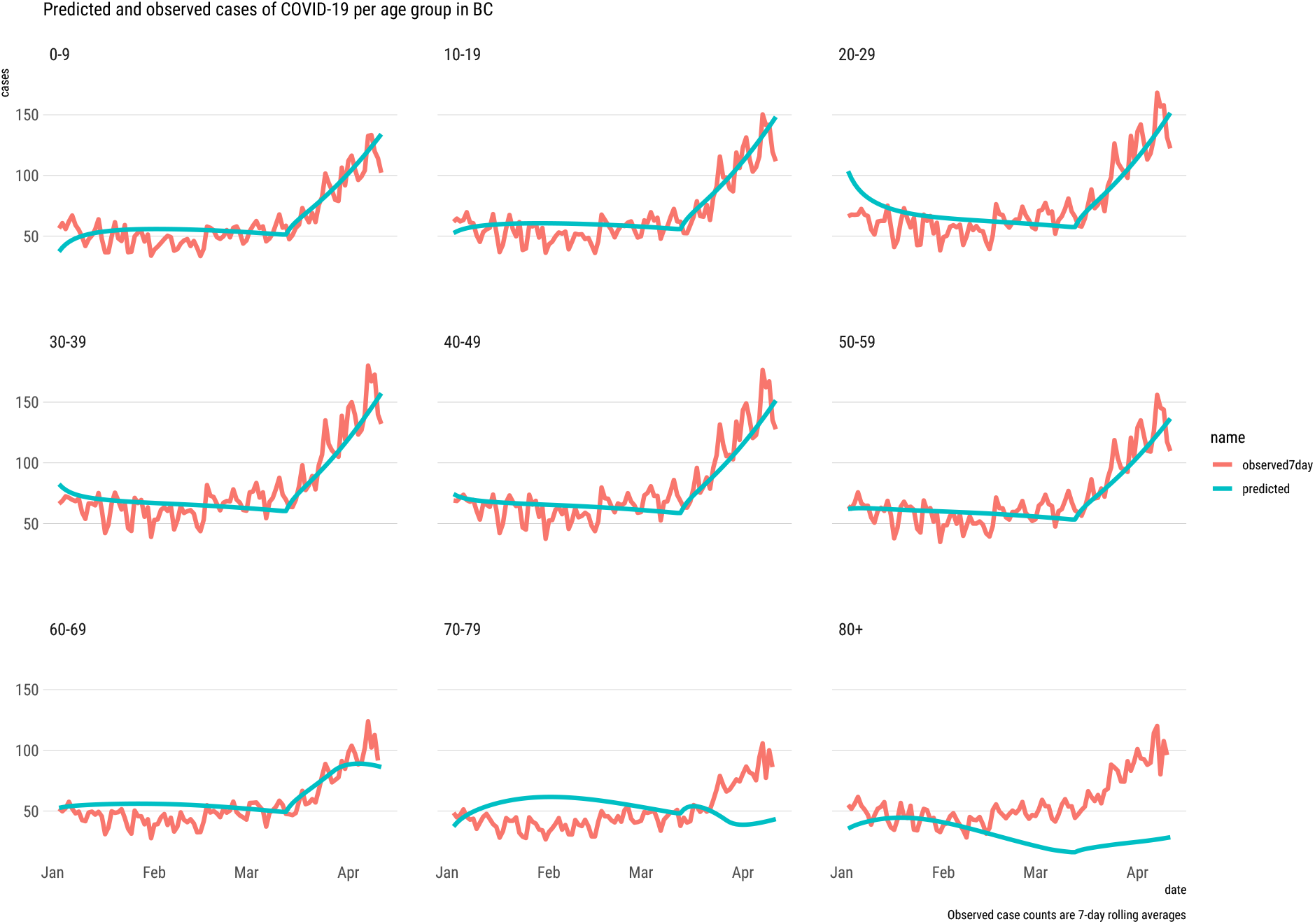
Observed and predicted case counts for different age groups when *v*_*p*_ = 0.90

**Figure B3.**
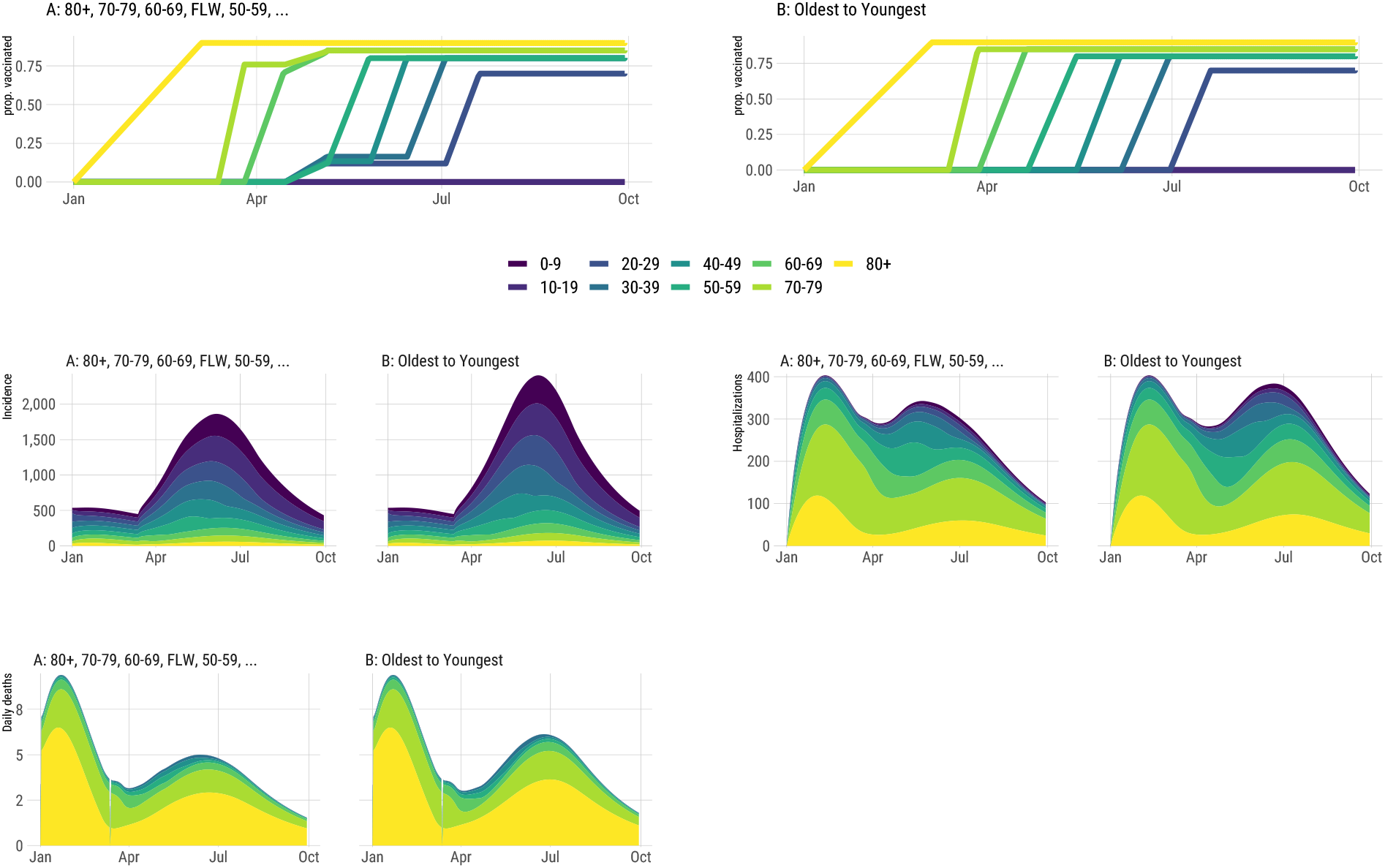
Top Panel: Projection of the progression of the vaccination for different age groups and front-line workers (FLW). Bottom Panel: Projection of COVID-19 cases, hospitalizations, and deaths from January 1st to October 1st, 2021 for *v*_*p*_ = 0.90

**Figure B4.**
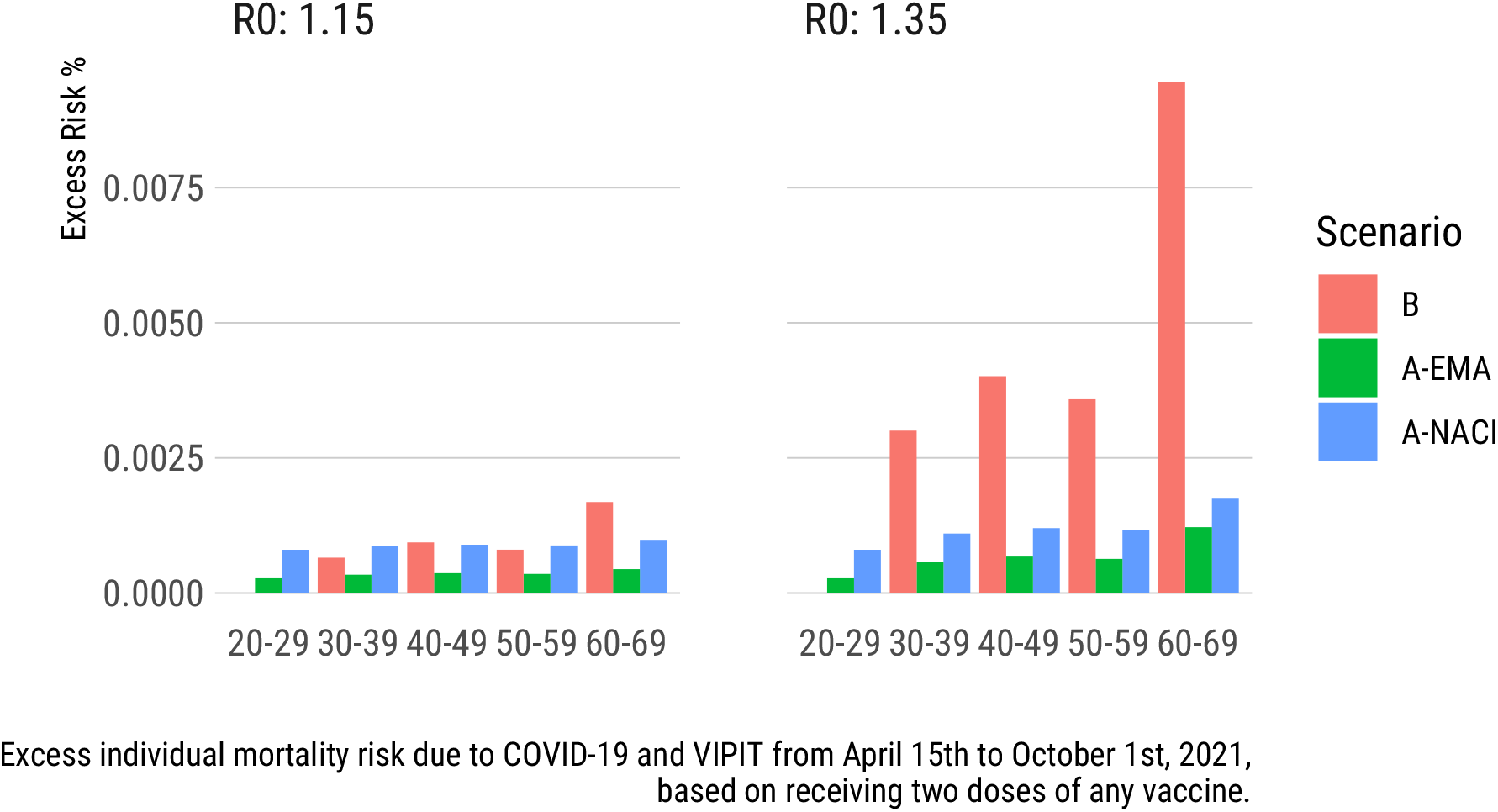
Comparison of excess mortality risk for different age groups based on the COVID-19 risk caused by delayed vaccination (B) and estimated residual COVID-19 and VIPIT risk (A) by the EMA and NACI when *v*_*p*_ = 0.90

**Figure B5.**
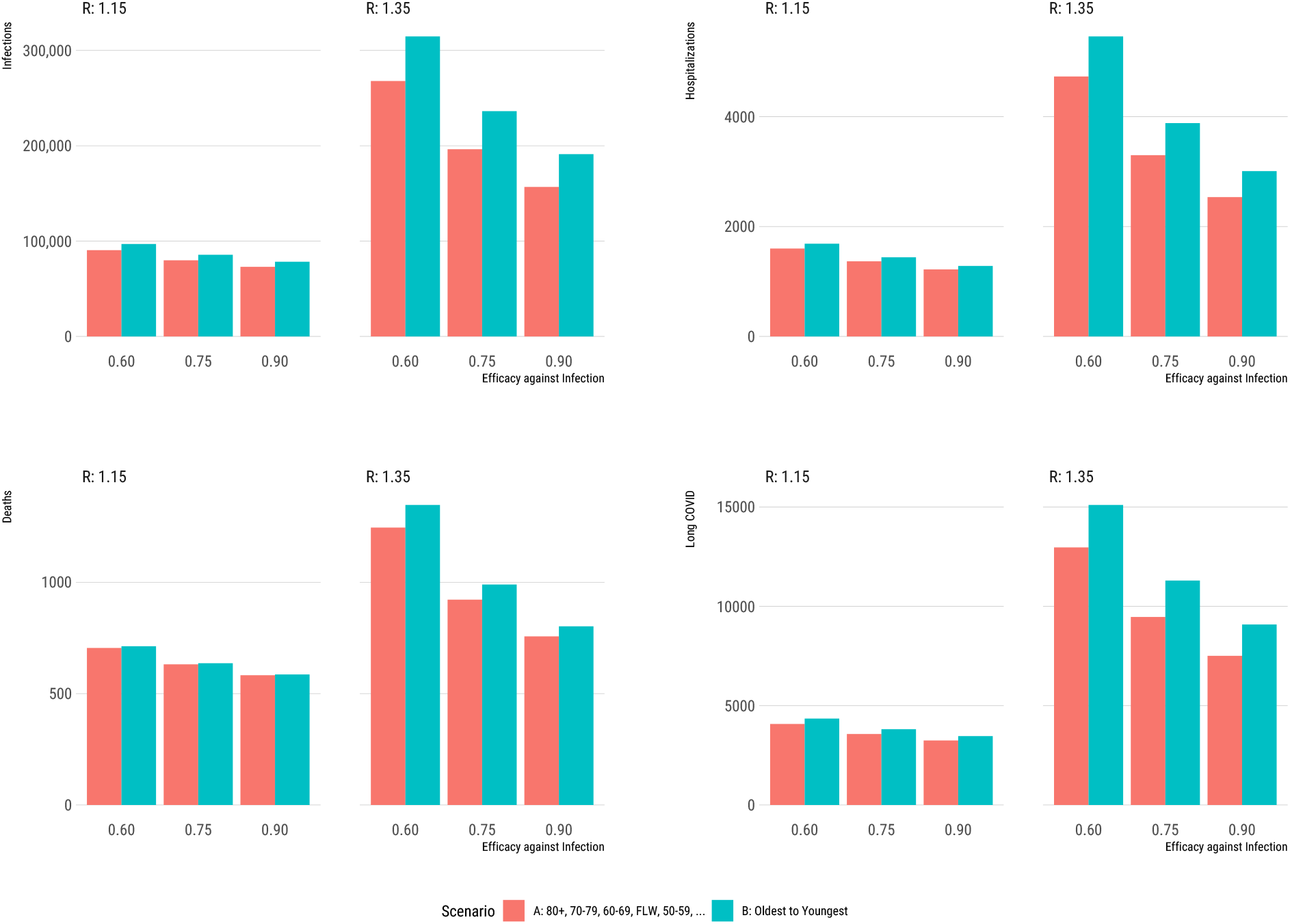
COVID-19 outcomes under different vaccination scenarios for different age groups and front-line workers (FLW) when *v*_*p*_ = 0.90

**Figure B6.**
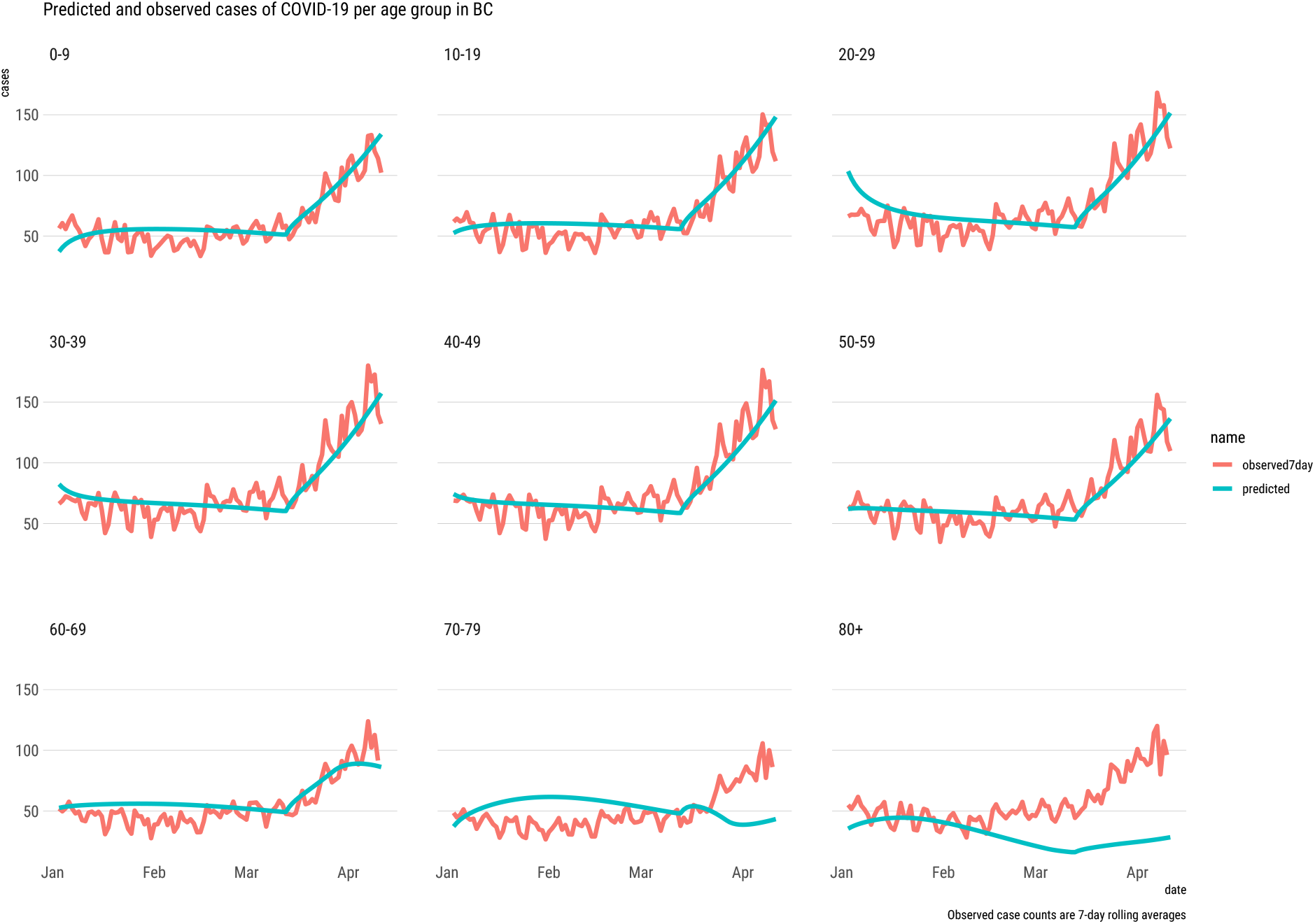
Observed and predicted case counts for different age groups when *v*_*p*_ = 0.60

**Figure B7.**
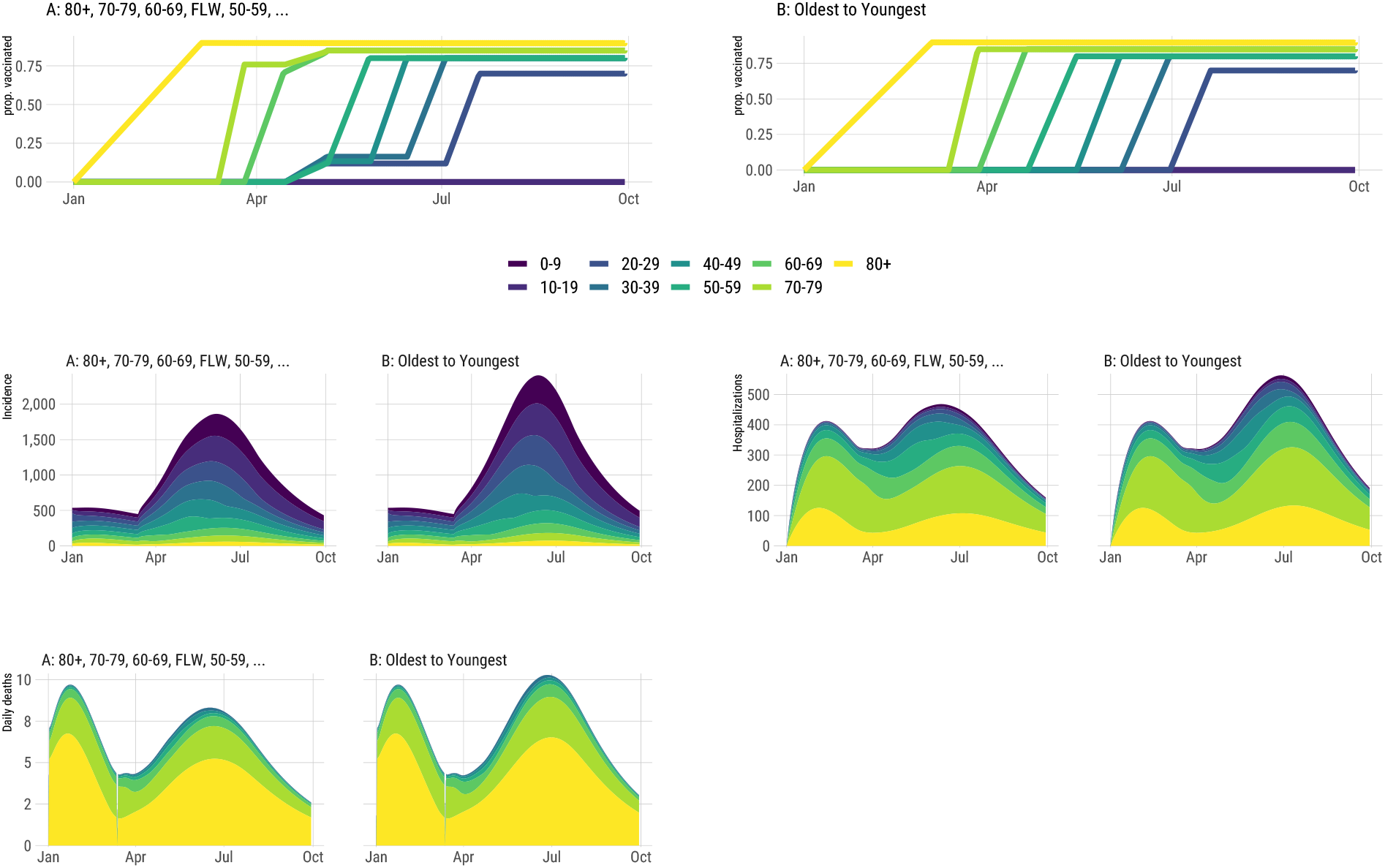
Top Panel: Projection of the progression of the vaccination for different age groups and front-line workers (FLW). Bottom Panel: Projection of COVID-19 cases, hospitalizations, and deaths from January 1st to October 1st, 2021 for *v*_*p*_ = 0.60

**Figure B8.**
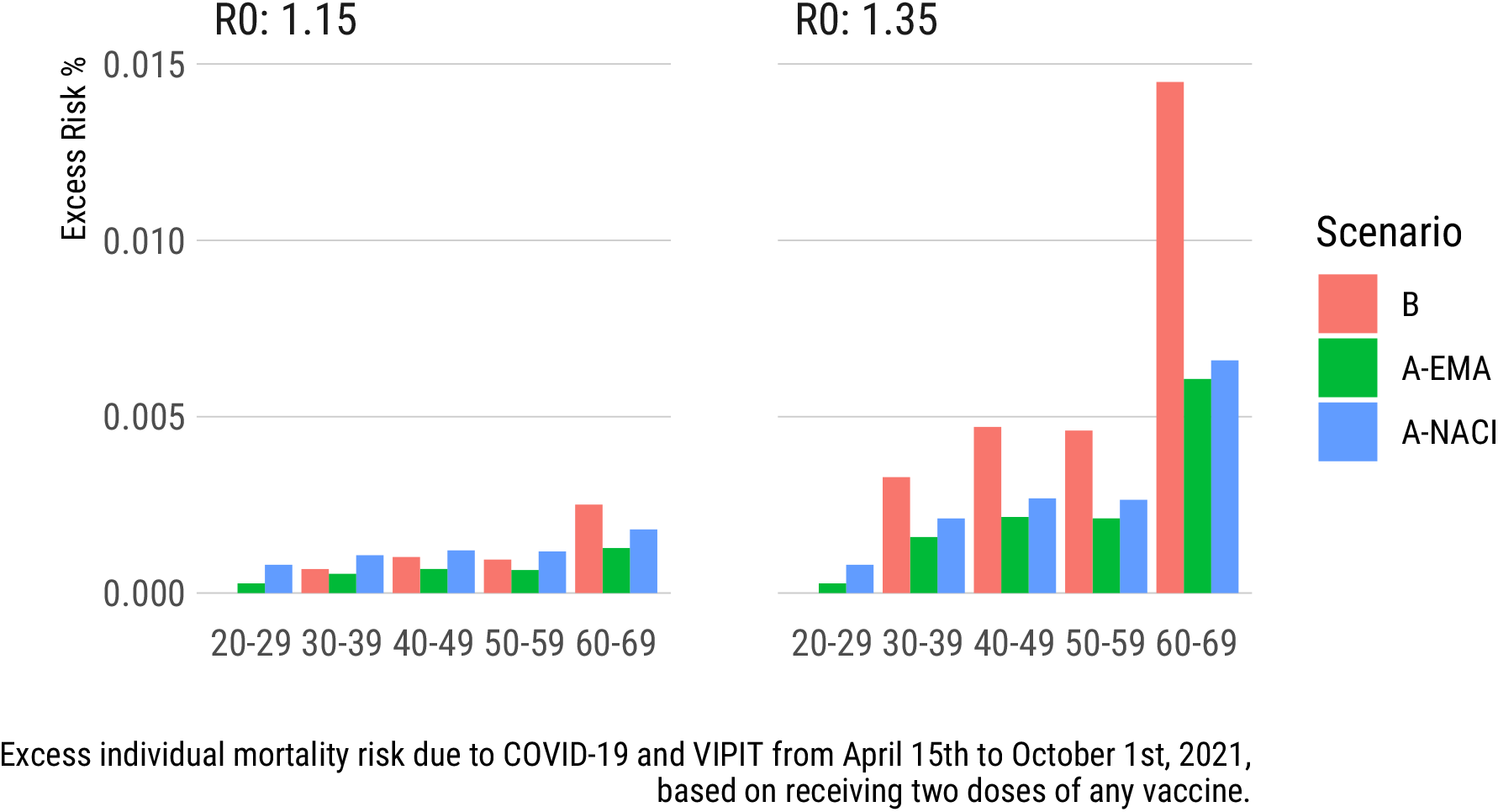
Comparison of excess mortality risk for different age groups based on the COVID-19 risk caused by delayed vaccination (B) and estimated residual COVID-19 and VIPIT risk (A) by the EMA and NACI when *v*_*p*_ = 0.90

**Figure B9.**
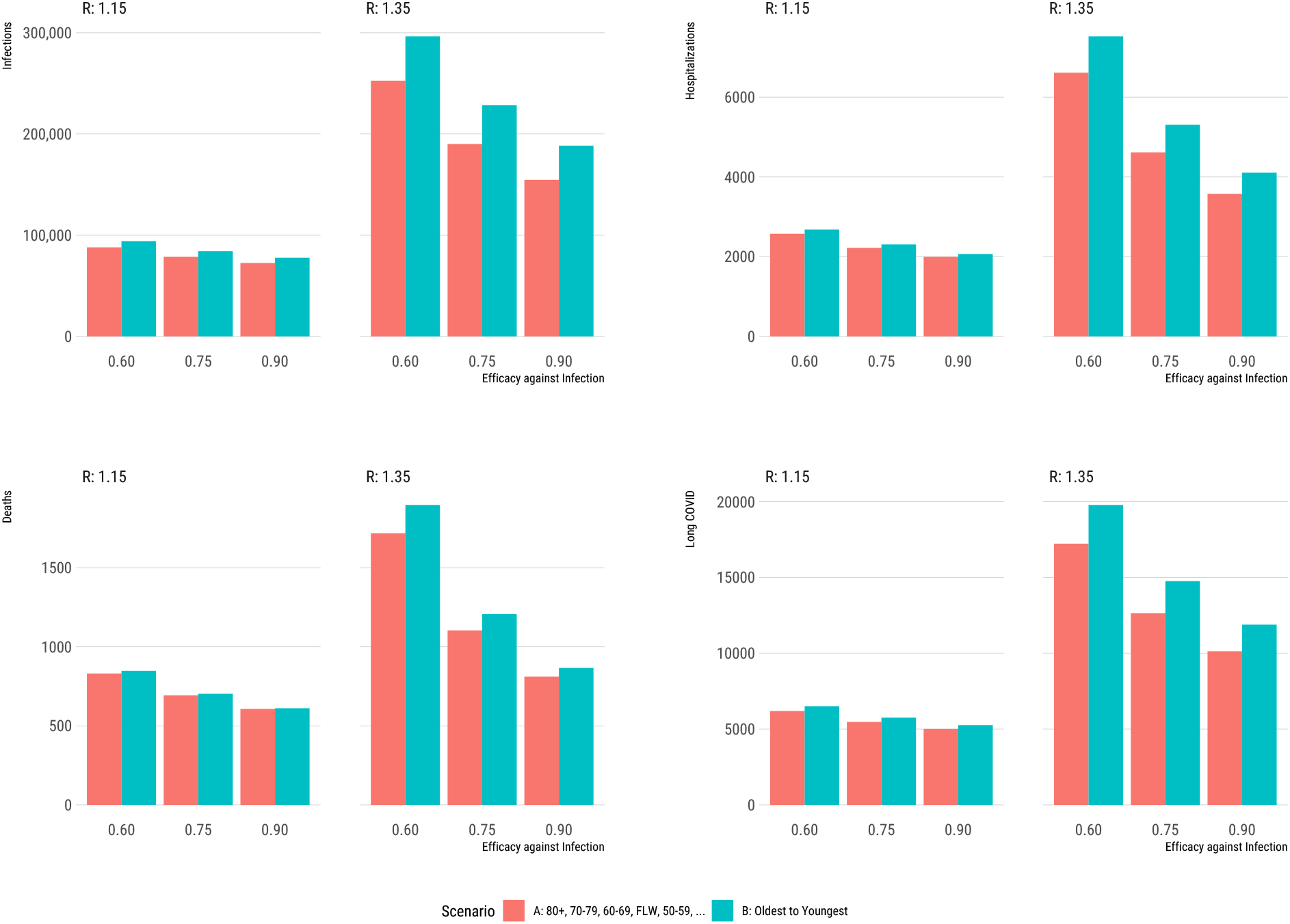
COVID-19 outcomes under different vaccination scenarios for different age groups and front-line workers (FLW) when *v*_*p*_ = 0.90

